# Combining pharmacogenetics and patient characteristic polygenic scores to improve outcome prediction for Calcium Channel Blocker treatment

**DOI:** 10.1101/2023.02.10.23285767

**Authors:** Deniz Türkmen, Jack Bowden, Jane A.H. Masoli, João Delgado, Chia-Ling Kuo, Luke C. Pilling, David Melzer

**Author notes:** **Corresponding Author** Professor David Melzer, College House (1.07), University of Exeter St. Luke’s Campus, Magdalen Road, Exeter, EX1 2LU.

## Abstract

**Background:** Calcium channel blockers (CCBs) are common antihypertensive medications. Pharmacogenetic variants affect CCB clinical outcomes, although effect sizes are modest in community samples. Variation in patient characteristics may also predict CCB outcomes, and variation attributable to relevant polygenic scores is less prone to confounding. We aimed to test associations between genetically predicted patient characteristics plus pharmacogenetic variants with CCB outcomes in a large community cohort.

**Methods:** We extended our analysis of 32,000 UK Biobank dihydropiridine CCBs treated participants (mean duration 5.9 years) testing 23 variants, where *NUMA1* rs10898815 and *RYR3* rs877087 showed the most robust associations (for discontinuation and heart failure, respectively). We calculated polygenic scores for systolic and diastolic blood pressures (SBP and DBP), body fat mass, waist hip ratio, lean mass, serum calcium, eGFR, lipoprotein A, urinary sodium, and liver fibrosis. Outcomes were CCB discontinuation, heart failure, coronary heart disease and chronic kidney disease.

**Results:** For heart failure, the highest risk 20% of polygenic scores for fat mass, lean mass and lipoprotein A were associated with increased risks (Hazard-Ratio (HR)Fat-mass 1.46, 95% CI 1.25-1.70, p=1*10-6; HRLean-mass 1.20, 95%CI 1.04-1.38, p=0.01; HRLipoproteinA 1.29, 95% CI 1.12 to 1.48, p= 4*10-4), versus the lowest risk 20% of each score respectively. Across the cohort, *RYR3* T-allele modestly increased heart failure risks (HR 1.13: 1.02-1.25) versus non-carriers, but in subsets with high fat mass, lean mass, and lipoprotein A scores, estimates were substantially larger, e.g., in females aged 65-70 the heart failure Relative Risk was 4.4 (95% CI 1.54-12.4) versus no T-alleles and low scores.

For CCB discontinuation, high polygenic scores for fat mass and lean mass increased risks versus the lowest 20%, whereas high SBP and DBP scores decreased discontinuation risks. Hazard ratios for discontinuation with the pharmacogenetic NUMA1 rs10898815 A-allele (overall HR 1.07: 1.02-1.12) were higher (HR 1.17: 1.05-1.29) in those with high polygenic scores for fat mass and lean mass.

**Conclusion:** Polygenic scores affecting adiposity and lipoprotein A levels add to known pharmacogenetic variants in predicting key clinical outcomes in CCB treatment. Combining pharmacogenetic variants and relevant individual characteristic polygenic scores may help for personalizing prescribing.

**What is needed, what do we add?:** We previously showed that pharmacogenetic variants in *RYR3* and *NUMA1* were associated with key clinical outcomes in community CCB patients, although effect sizes were modest. Various patient characteristics reportedly affect CCB outcomes. We therefore tested effects of relevant patient characteristics using polygenic scores. They minimize the effect of unmeasured confounders as genotypes are invariant since conception and reflect lifetime exposure to the risk factor. We showed that combining associated scores with the pharmacogenetic variants improved outcome prediction.

## 1 Introduction

High blood pressure (BP) is a major modifiable factor affecting cardiovascular disease morbidity and mortality. Dihydropiridine calcium channel blockers (dCCB, e.g., amlodipine) are among the most commonly prescribed first-line treatments for hypertension, recommended by major international guidelines^1,2^.

Pharmacogenetics is a subset of personalized medicine studying how genetic variation affects drug response and adverse events^3^. Several pharmacogenetic variants are reported in The Pharmacogenomics Knowledge Base (PharmGKB, searched January 2023) ^4,5^ to influence CCB response with modest levels of clinical evidence (levels 3-4, where level 1 out of 4 is the best evidence). In our previous work in the UK Biobank cohort of 32,000 patients receiving dCCB medications in routine clinical care, we showed that alleles in genes *NUMA1, RYR3, CYP3A5, ADRA1A* and *APCDD1* increased risks for adverse outcomes such as heart failure and coronary heart disease, and for discontinuation for dCCBs^6^. However, the effects sizes were modest, for example the Hazard Ratio for heart failure in *RYR3* rs877087 T allele carriers in dCCB patients was 1.13 (95% CI 1.02-1.25), compared to participants with no T alleles. Yet we observed no association in participants not receiving dCCBs, demonstrating the pharmacogenetic effect of *RYR3* rs877087 T allele on heart failure in dCCB prescribing.

Individual patient characteristics, including age, weight, renal, and hepatic functions, may also modify response to dCCBs, as these medications are lipophilic with high protein-binding capacity, hepatic metabolism and renal excretion^7,8^. Previously reported individual characteristics affecting dCCB response include weight, adiposity, baseline blood pressure, biological biomarkers (including serum calcium and urinary sodium), renal and hepatic functions, and lipoprotein ^9–13^ but the links remain inconclusive^14^. This is likely due to differences in risk factors studied, low power due to small sample sizes, a focus on selected patient groups not necessarily representative of clinical practice, and biases common to observational study designs, including confounding and reverse causation.

Much work is ongoing to integrate pharmacogenetic information to personalized prescribing - optimizing treatment effectiveness and reducing side-effects^3,15–19^. To date there has been limited discussion of whether genetically determined individual patient characteristics (using polygenic scores) could be useful predictors of drug response^20^. Polygenic scores reflect individuals’s genetic predisposition for a phenotype, derived by summing the number of trait-increasing alleles they carry and weighting each variant’s contribution according to its effect size. Analyzing polygenic scores can minimize the effect of unmeasured confounders as genetic variants are fixed at conception and reflect lifetime exposure to the risk factor^21^. Polygenic scores are emerging as important tools for personalized medicine^17^ and have shown utility in identifying individuals with e.g. exceptionally high genetic risks for CHD^22^. In addition to individual prediction, polygenic scores have been used to model the effects of risk traits, including BMI^23^ for hypertension and CHD, and systolic and diastolic blood pressure genetic risk for phenotypic hypertension or diagnose cardiovascular diseases^24,25,26^. A recent UK Biobank study examining BP and low-density lipoprotein polygenic scores found that, among those treated for hypertension, an increased SBP polygenic score was still associated with uncontrolled hypertension^27^. However, there have been few studies utilizing polygenic scores and pharmacogenetic variants to predict clinical outcomes.

As noted above, individual patient characteristics may influence clinical outcomes during CCB treatment, alongside pharmacogenetic variation. Here, we aimed to test whether polygenic scores for genetically predicted patient characteristics are associated with relevant clinical outcomes independent of pharmacogenetic variants. We also aimed to test whether combining polygenic score risks with previously reported pharmacogenetic risk factors improved prediction of the selected clinical outcomes, in a large cohort of 32,000 UK Biobank community participants prescribed dCCBs in routine clinical care.

## Methods

### Cohort descriptions

#### UK Biobank cohort

503,325 community-based volunteers aged 40-70 years were recruited in UK Biobank (UKB). Assessments were done in one of 22 assessment centers in Wales, Scotland, or England in 2006-2010 ^28^. Lifestyles and health information, as well as blood samples for biochemical and genetics analyses were gathered. Linked GP (General Practice) data are available for 230,096 participants from between Jan 1990 and August 2017.

#### General practice (GP) Data

UKB included more than 57 million prescriptions for 230,096 (45.7%) participants from the linked GP data that is available up to 31 May 2016 (England TPP system) and 31 August 2017 (Wales EMIS/Vision system). We identified prescribing information for dCCB medications (amlodipine, felodipine, lacidipine, lercanidipine, nimodipine, nisoldipine, nifedipine, nitrendipine and nicardipine) and dates of prescriptions, using drug codes available (in clinical Read v2, British National Formulary (BNF), or dm+d (Dictionary of Medicines and Devices) format, depending on suppler. The UK National Institute for Health and Care Excellence (NICE) BNF database (https://bnf.nice.org.uk) was our primary source to detect medication and brand names prescribed in the NHS that met our search criteria. We analyzed the dihydropyridine subset of CCBs together because effect sizes for the variants against the outcomes were consistent, although with reduced statistical significance due to reduced sample size (n= 31,357 amlodipine and n= 6,854 others, vs. n= 43,891 total dCCB), for details see the previous analysis^6^.

We ascertained cardiovascular events from hospital admissions inpatient records with up to 14 years follow-up after baseline assessment (HES in England up to 30 September 2021: data from Scotland and Wales censored to 31 August 2020 and 28 Feb 2018, respectively), covering the period up to the date of censoring of primary care prescribing data. Diagnoses of incident heart failure, coronary heart disease (myocardial infarction/angina) and chronic kidney diseases were ascertained using ICD-10 codes (described previously^6^).

Discontinuation was defined as patients having a date of last prescription at least 90 days prior to the censoring date which is either the date of deduction (removal from GP list, where available) or 28.02.2016 where no deduction date existed. Depending on primary care provider, data after 28.02.2016 was often incomplete (See UK Biobank resource 591^29^).

#### Genotype data

Primary analysis included 451,367 participants (93% of 481,000 with genotype data available) identified as ancestrally European (determined by genetic clustering, as explained previously ^30^): sample sizes from other genetic backgrounds were deemed too small to examine separately. Genotyping in UK Biobank is from Affymetrix microarrays (800,000 directly genotyped variants) plus imputation (Haplotype Reference Consortium) has been described in detail previously ^6,29^.

#### Polygenic score

We calculated polygenic scores in UK Biobank participants by summing the number of studied risk factors-increasing alleles carried, multiplied by the known effect size of the allele on the risk factor levels using genetic instruments from publicly available large-scale genome wide association studies (GWAS) from the Open GWAS platform^31^ – see Table 1 for list. We estimated associations between polygenic scores for patients’ characteristics including systolic BP, diastolic BP, whole body fat mass, appendicular lean mass, waist-hip ratio, serum calcium, lipoprotein A, sodium, estimated glomerular filtration rate (eGFR), and fibrosis, with outcomes such as heart failure and discontinuation.

**Table 1.**
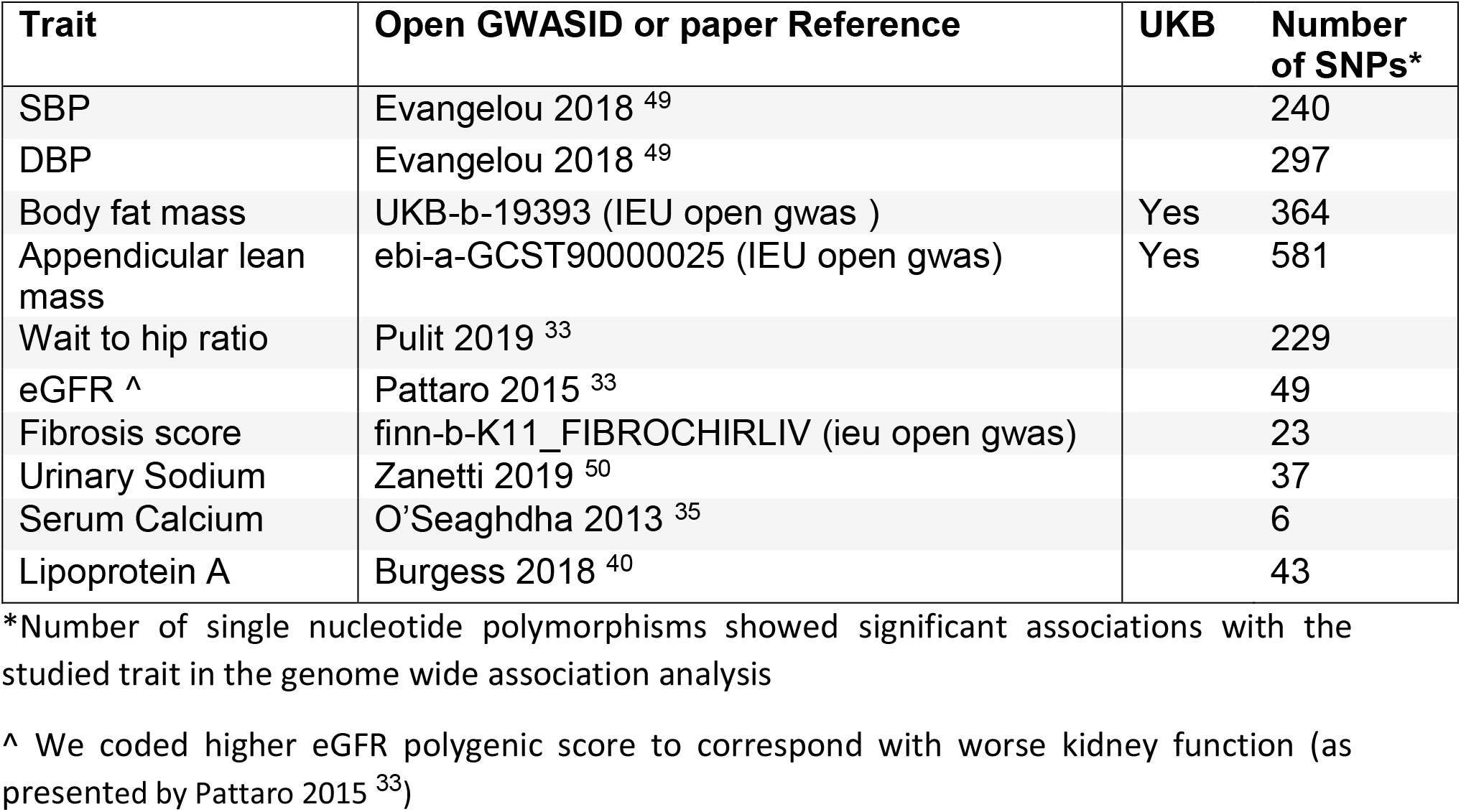
List of traits and GWASs used in polygenic scores.

| Trait | Open GWASID or paper Reference | UKB | Number of SNPs* |
| --- | --- | --- | --- |
| SBP | Evangelou 2018 <sup>49</sup> |  | 240 |
| DBP | Evangelou 2018 <sup>49</sup> |  | 297 |
| Body fat mass | UKB-b-19393 (IEU open gwas ) | Yes | 364 |
| Appendicular lean mass | ebi-a-GCST90000025 (IEU open gwas) | Yes | 581 |
| Wait to hip ratio | Pulit 2019 <sup>33</sup> |  | 229 |
| eGFR ^ | Pattaro 2015 <sup>33</sup> |  | 49 |
| Fibrosis score | finn-b-K11_FIBROCHIRLIV (ieu open gwas) |  | 23 |
| Urinary Sodium | Zanetti 2019 <sup>50</sup> |  | 37 |
| Serum Calcium | O'Seaghdha 2013 <sup>35</sup> |  | 6 |
| Lipoprotein A | Burgess 2018 <sup>40</sup> |  | 43 |

\*Number of single nucleotide polymorphisms showed significant associations with the studied trait in the genome wide association analysis
^ We coded higher eGFR polygenic score to correspond with worse kidney function (as presented by Pattaro 2015 <sup>33</sup>)

In mathematical notation the polygenic score variable *Ŝ* is derived as:

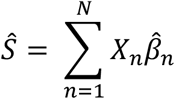

where *X*_*n*_ is the number of trait-increasing alleles of variant *n* weighted by their published effect size estimate 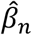, for each of the *N* genetic variants (*n* = 1 … *N*).

#### Primary Analysis: Survival Analysis

Age at first prescription was categorized as “40-44”, “45-59”, “50-54”, “55-59”, “60-64”, “65-69”, “70-74” and “75-80”.

We aimed to extend our previous pharmacogenetics study in 32,000 UKB dCCB patients, adding the effects of polygenic scores of patients’ characteristics on the same outcomes studied. In our pharmacogenetic analyses^6^, the most striking results were for *NUMA1* rs10898815 increasing the risk of treatment switch (significant after Benjamini–Hochberg adjustment for multiple statistical testing - adjusted p = 0.04) and *RYR3* rs877087 increasing the risk of heart failure (plausibility of the associations increase by the burden of prior evidence).

We used Cox proportional hazards regression models adjusted for sex and age at first prescription. The primary outcomes were incident heart failure diagnoses and discontinuation of dCCB prescribing. We opted to use discontinuation over ‘switching antihypertensive treatment’ to better capture patients who are no longer prescribed dCCBs for any reason. Secondary outcomes were coronary heart and chronic kidney disease. Patients were included in the analyses if they had at least 2 dCCB prescriptions in a year and were older than age 40 at the first prescription (Details are described elsewehere^6^). Patients entered the model on the date of the first prescription and exited with the censoring date described above in the discontinuation model.

We first conducted analyses with continuous polygenic scores, and then repeated the analyses with the 5 quintiles of polygenic scores to allow for non-linearity. For the models using continuous polygenic scores, we standardized the scores using the z-transform to be able to compare the hazard ratios across different units.

STATA (v15.1) software and R (v4.2.1) were used for the analyses. ‘stset’, ‘stcox’ commands in STATA, and ‘coxph’ from the ‘survival’ package (v3.4-0) in R was used to fit Cox proportional hazards models.

#### Secondary Analysis: Combining Pharmacogenetics and Polygenic scores of risk factors

We used ‘predict’ command in R to get individual level predictive models for discontinuation and heart failure risks, to see if the modest effect sizes of pharmacogenetic variants increase in certain patients with significant high polygenic scores. The ‘predict’ command compares models with the minimum risk group (non-carriers of the allele and having the low genetic risk). To prevent sex and age-related bias, we examined female and male patients separately across age groups. First model was with the carriers of the allele, and having bottom 20% of the polygenic score vs compared to the minimum risk group, and the second model was with the carriers of the allele, and having top 20% of the polygenic score vs compared to the minimum risk group, non-carriers of the allele and with bottom 20% of polygenic scores (e.g., females aged 65-70 with low genetic risk).

##### Heart failure

Carrying at least one rs877087 T allele in *RYR3* (prevalence of 46% in UKB) increased the risk of HF with the HR 1.13 (95% CI 1.02-1.25) compared to non-carriers in UKB dCCB patients previously. Details can be found here^6^. The models included either op 20% or bottom 20% of the polygenic scores with significant result (body fat mass, lean mass and lipoprotein A)

##### Discontinuation

Carriers of rs10898815 GA allele (HR=1.10, 95 % CI 1.01 to 1.21) and AA allele in *NUMA1* (HR=1.18, 95% CI 1.07 to 1.31), and rs776746 TT allele in *CYP3A5* (HR 1.87, 95% CI 1.26 to 2.78) increased the risk of discontinuing dCCBs compared to their common homozygotes. We only took rs10898815 into account here due to the low prevalence of rs776746 TT (0.5% in this cohort). The same method was used here for individual level risk predications as above, examining for body fat mass and lean mass.

We previously observed dominant effects for rs877087 T allele and rs10898815 A allele, so we modelled the carriers (heterozygotes plus homozygotes) compared to non-carriers (homozygous reference) throughout.

### Sensitivity analyses

We conducted 3 sensitivity analyses in the pharmacogenetic analyses; assessed for additional antihypertensions within the dCCB prescription time, analysis of amlodipine only and other dCCBs, and analysis of unrelated participants only. The results of the analyses remained consistent with the main analysis; therefore, we did not present the sensitivity analysis results here.

#### 1 Two-sample MR methods

To test whether associations between genetically predicted body fat mass and dCCB clinical outcomes (HF, CHD and discontinuation) – the primary results – were potentially biased, we used summary statistics (or “two sample”) MR methods, to exploit the advantages of the different approaches. We used R package ‘TwoSampleMR‘ (v0.5.6) to estimate the inverse-variance weighted (IVW) effect of each individual exposure-associated variant on the outcome – akin to the estimate from the polygenic scores. The IVW (and polygenic score approach) assumes there is balanced horizontal pleiotropy (i.e. with a zero mean); we therefore additionally applied ‘weighted median’ (assumes less than 50% of the weight in the analysis comes from invalid instruments) and ‘MR-Egger’ (allows for unbalanced pleiotropy providing genetic variants’ effect on body fat mass is not correlated with their pleiotropic effects on the outcome) approaches to test robustness of the estimate. The MR-Egger method additionally estimates an intercept and deviation from the null is used to test for possible unbalanced pleiotropy. We also calculated the 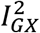 statistic to check for bias due to the ‘NO Measurement Error’ (NOME) assumption (i.e., where the SNP-exposure association is assumed to be known, rather than estimated). Finally, ‘leave one out’ analysis tests whether the result is overly influenced by a single variant.

#### 2 Excluding existing heart failure diagnoses

NICE report that amlodipine, nifedipine, felodipine and nicardipine do not cause deterioration in heart failure^32^. This advice could mean that more severe patients might have been prescribed dCCBs. In order to minimize the potential for selection bias, we repeated the model for heart failure excluding patients with existing heart failure diagnoses prior to the dCCB prescriptions.

## 2 Results

There were 32,360 (45.6% female) study participants with prescribed dCCB in primary care records. The mean age at first dCCB prescription was 61.3 (SD 7.7) years and the median numbers of prescriptions per year was 8.2 (interquartile range (IQR) 6.6 to 13, range 2 to 25). The mean prescription period was 5.9 (SD 5.2) years, the median was 4.4 (IQR 1.6 to 9.1). See Table 2 for patient characteristics.

**Table 2.** Characteristics of UKB patients prescribed dCCB.

| **GP prescribed** |  |
| --- | --- |
| Number of participants | 32,360 |
| Females, n (%) | 17,590(45,6) |
| Age at first prescription |  |
| Minimum: maximum | 40:79.3 |
| Mean (SD) | 61.3(7.7) |
| Number of prescriptions in a year |  |
| Minimum: maximum | 2:25 |
| Mean (SD) | 9.6(4.4) |
| Years between first and last prescription |  |
| Minimum: maximum | 0.08:39.9 |
| Mean (SD) | 5.9(5.2) |
| MI/angina^^ pre prescription of dCCBs | 3,026(7.7) |
| CKD^^ pre prescriptions of dCCBs | 229(0.6) |
| Heart failure^^ pre prescriptions of dCCBs | 333(0.8) |
| Prescribed other antihypertensive during Dccb | 23,971(61) |
| Discontinue dCCBs | 10,095(31.4) |
| MI/angina^^ post prescription of dCCBs | 7,430 (18.9) |
| CKD^^ post prescriptions of dCCBs | 2,940 (7.5) |
| Heart failure^^ post prescriptions of dCCBs | 2,292(5.8) |

*European-ancestry participants with >1 dCCB prescription in the available GP prescribing data.*
*CKD, chronic kidney disease;*
*dCCB, dihydropyridine calcium-channel blocker.*
*^^Hospital diagnosed diseased*

### 1 Polygenic scores

Of the ten polygenic scores tested, four were associated with increased risk of a studied CCB adverse event (Figure 1): body fat mass, lean mass, lipoprotein A, and eGFR and two were negatively associated: SBP and DBP.

**Figure 1.**
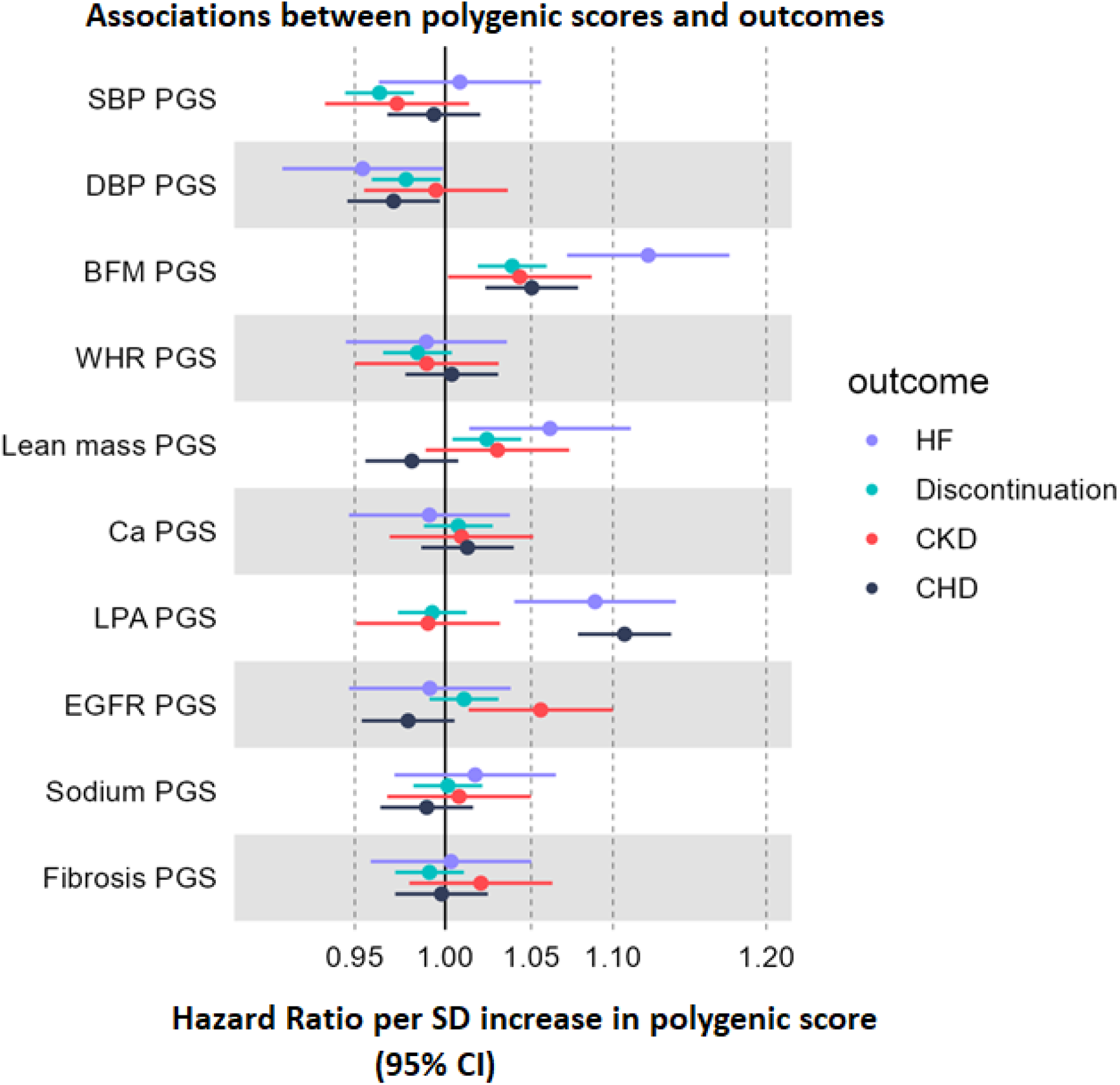
Forest plot of associations between patients’ characteristics polygenic scores and the risks of increased dCCB adverse events and discontinuation in 32,000 UK Biobank patients.

#### Body fat mass polygenic score

Body fat mass polygenic score was associated with increased risk of dCCB discontinuation (HR per SD increase in polygenic score 1.04: 95% CI 1.02-1.06, *p*=1.3×10^−4^), of heart failure (HR 1.12: 95% CI 1.07-1.18, *p=* 9.6×10^−7^), and of coronary heart disease (HR 1.05: 95% CI 1.02-1.08, *p*=2.5×10^−4^) in the Cox proportional-hazards model. For the details see supplementary table 1.

As polygenic score associations are often non-linear, we also stratified patients into 5 quintiles (Figure 2): those in the highest risk 20% of body fat mass polygenic score had increased risk for heart failure by 46% (HR=1.46, 95% CI 1.25 to 1.70, *p*=1*10^−6^) compared to the lowest 20%. The difference in measured body fat between the top and bottom quintiles was 6 kg (95%CI 5.7-6.3) in the linear regression model adjusted for age, sex and the top 10 genetic principal components. They also had increased risk for discontinuation of dCCBs (HR 1.13, 95% CIs 1.06 to 1.20, *p*=2.×10^−4^) and coronary heart disease (HR 1.14, 95% CI 1.05-1.24, p=0.003) (Table 3). Additional results are presented in supplementary table 2.

**Table 3.** Cox Proportional-hazards models between body fat mass 20% quintiles and outcomes in patients treated with dCCB.

| Outcome | Body fat mass polygenic score group | n | % n in N | HR | 0.95 | CI | p |
| --- | --- | --- | --- | --- | --- | --- | --- |
| **Heart Failure**<br>(n=1,838/N=31,051) | 1 | 262 | 4.7 | Ref | Ref | Ref | Ref |
| | 2 | 325 | 5.5 | 1.20 | 1.02 | 1.41 | $3 \times 10^{-2}$ |
| | 3 | 385 | 6.1 | 1.31 | 1.12 | 1.53 | $8 \times 10^{-4}$ |
| | 4 | 393 | 6.1 | 1.30 | 1.12 | 1.53 | $9 \times 10^{-4}$ |
| | 5 | 473 | 7 | 1.46 | 1.26 | 1.70 | $8 \times 10^{-7}$ |
| **Discontinuation**<br>(n=10,095/N=32,120) | 1 | 1,697 | 29.3 | Ref | Ref | Ref | Ref |
|  | 2 | 1,842 | 30.4 | 1.05 | 0.98 | 1.12 | 0.17 |
| | 3 | 2,104 | 32.4 | 1.12 | 1.05 | 1.19 | $7.5 \times 10^{-4}$ |
| | 4 | 2,121 | 32 | 1.09 | 1.02 | 1.16 | $7.3 \times 10^{-3}$ |
| | 5 | 2,331 | 33.2 | 1.13 | 1.06 | 1.2 | $2 \times 10^{-4}$ |
| **Coronary Heart Disease**<br>(n=5555/N=30,722) | 1 | 931 | 16.7 | Ref | Ref | Ref | Ref |
|  | 2 | 1,004 | 17.2 | 1.05 | 0.96 | 1.14 | 0.3 |
| | 3 | 1,131 | 18.2 | 1.1 | 1.01 | 1.2 | $2.6 \times 10^{-2}$ |
| | 4 | 1,210 | 19 | 1.16 | 1.06 | 1.26 | $8.4 \times 10^{-4}$ |
| | 5 | 1,279 | 19 | 1.14 | 1.05 | 1.24 | $2.6 \times 10^{-3}$ |

*n=number of events*
*N=total number of patients*

**Figure 2.**
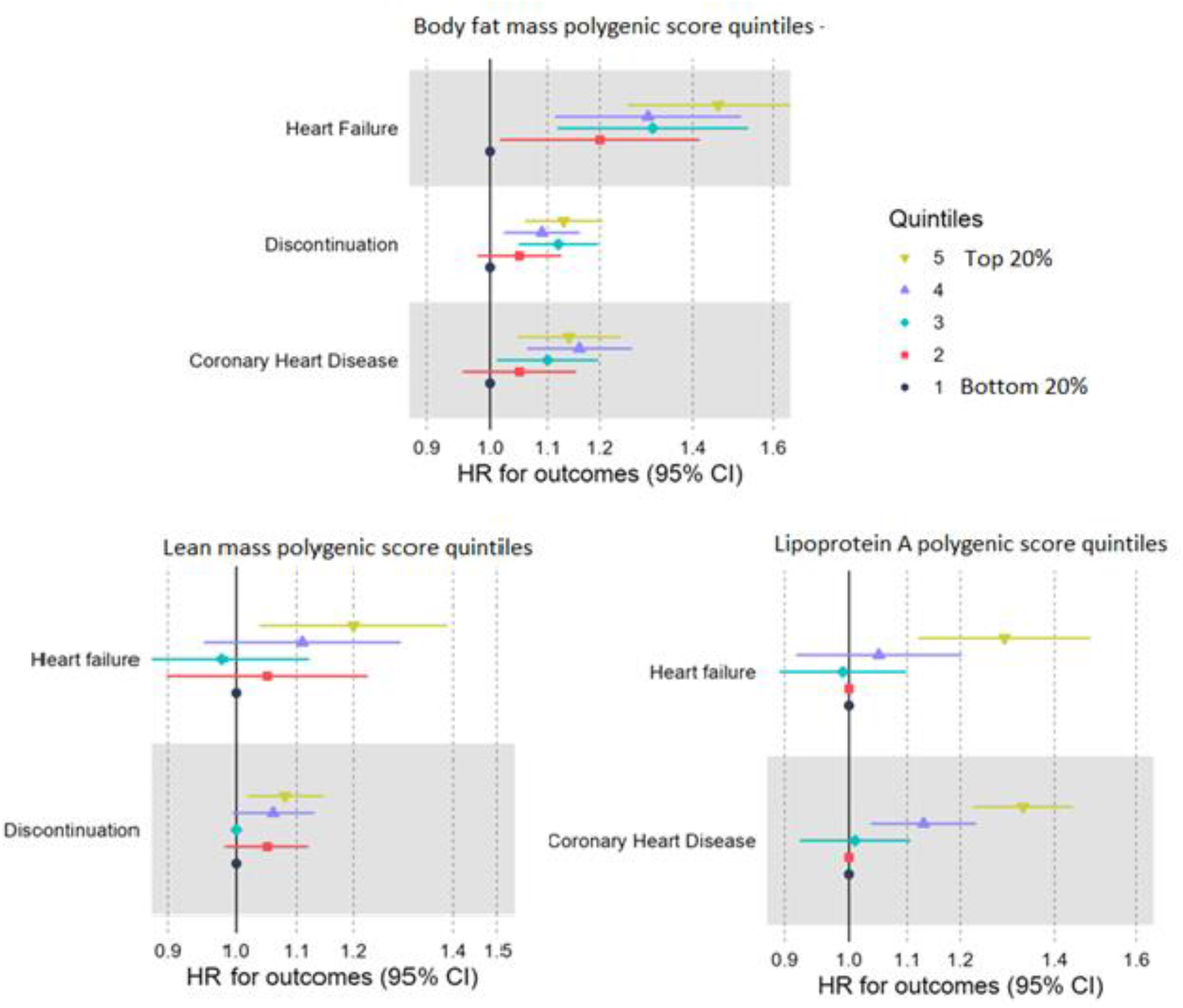
Forest plot of the associations between body fat mass, lean mass and lipoprotein A quintiles and outcomes.

#### Lean mass polygenic score

A higher lean mass polygenic score was associated with increased risk of heart failure (HR per SD increase in polygenic score 1.06: 95% CI 1.01-1.11, p=0.01) and discontinuation (HR 1.02, 95% CI 1-1.04, p=0.02)

When patients were stratified into 5 quintiles, patients at the highest risk 20% of lean mass polygenic score had increased risks of heart failure and discontinuation compared to the bottom 20% by 20% (HR 1.2, 95% CI 1.04 to 1.38, *p*=0.01) and by 8% (HR 1.08, 95% CI 1.02 to 1.15, *p*=0.01, respectively) (Figure 2).

#### Lipoprotein A polygenic score

A higher lipoprotein A polygenic score was associated with increased risk for heart failure (HF) and coronary heart disease (CHD) (HR_HF_ per SD increase in polygenic score 1.09, 95% CI 1.04-1.14, p=2.7×10^−4^ and HR_CHD_ per SD increase in polygenic score 1.11, 95% CI 1.08-1.14, p=4.5×10^−14^).

When patients were stratified into 5 quintiles, patients with the highest risk 20% lipoprotein A score had an increased risk of HF by 29% (HR 1.29, 95% CI 1.12 to 1.48, *p=* 0.0004) and CHD by 33% (HR 1.33, 95%CI 1.22 to 1.44, *p=*0.8×10^−11^) compared to the lowest 20% despite the dCCB treatment (Figure 2).

#### Systolic BP and Diastolic BP polygenic scores

Higher systolic BP and diastolic BP scores were associated with lower discontinuation rates (HR_SBP_ per SD increase in polygenic score 0.96, 95% CI 0.94-0.98 and HR_DBP_ per SD increase in polygenic score 0.98, 95% CI 0.96-0.99) (Figure 1 and supplementary table 1).

Patients at the highest 20% burden of SBP or DBP polygenic score had lower discontinuation rates, HR _SBP=_ 0.93 (95% CI 0.88 to 0.99) and HR _DBP_=0.93 (95% CI 0.88 to 0.99) compared to the lowest 20%. Those patients with highest burden of DBP polygenic score also were less likely to have heart failure and coronary heart disease compared to the lowest 20% (HR 0.86, 95% CI 0.74 to 0.99, *p*=0.04 and HR 0.91, 95% CI 0.84-0.99, p=0.03, respectively).

#### Estimated Glomerular Filtration Rate polygenic score

For the eGFR polygenic score, a higher score corresponds with lower measured eGFR (as presented by Pattaro 2015 ^33^). In our analysis of dCCB treated patients, higher eGFR polygenic score (I.e., worse function) was associated with increased risk for CKD, with the highest 20% having 1.19 (95% CI 1.04 to 1.36, *p*=0.01) times the risk versus the lowest (See supplementary table 1 for details)

### 2 Pharmacogenetics and Polygenic scores

Pharmacogenetic variants were independent predictors of adverse outcomes in combined analysis with the above polygenic scores.

In a Cox proportional hazards regression model for incident heart failure after initiating dCCB treatment, rs877087 T allele and polygenic scores for body fat mass, lean mass, and lipoprotein A had significant, independent effects (HR_rs877087_ 1.13, p=0.02; HR_body fat mass_ 1.08, p= 1.7×10^−6^; HR_lean mass_ 1.04, p=0.02, HR_Lipoprotein A_ 1.06, p=2×10^−4^), after adjusting for age at treatment initiation, sex and the top 10 genetic principal components. Across the whole cohort, carriers of the *RYR3* rs877087 T-allele had modestly increased heart failure risk (HR 1.13: 1.02-1.25). In the subset with the highest fat mass, lean mass, and lipoprotein A polygenic scores, the estimate was larger (HR 1.62, 95% CI 1.01-2.59); with the low polygenic scores, the estimate was not statistically significant (HR 0.92, 95% CI 0.49-1.73). (Supplementary Figure 1). The heart failure risk increased from 6.1% to 8.8% in the presence of high polygenic scores in T-allele carries patients.

For discontinuation, rs10898815 A allele carrier status and polygenic scores for body fat mass and lean mass also had significant, independent effects (HR_rs10898815_ 1.07, p=0.002; HR_body fat mass_ 1.03, p=10^−4^; HR_lean mass_ 1.01, p=0.04) and rates of discontinuation increased from 32% to 35% with the addition of highest polygenic scores of body fat mass and lean mass information (Supplementary Figure 2).

#### Risk Prediction across groups combining Pharmacogenetics and Polygenic Scores

In order to show the top risk group, we calculated 2 risk predictions for outcomes across age and sex groups compared to the group with minimum risk: group i carried a pharmacogenetic variant but had bottom 20% polygenic scores for the significant characteristics, and group ii carried a pharmacogenetic variant and top 20% polygenic scores.

#### Heart failure

Male dCCB patients were 1.8 times more likely to develop HF compared to female dCCB patients adjusting for age at first prescription, 10 genetic principal components and assessment center (HR=1.8, 95% CI 1.6 to 2).

The relative risk of incident heart failure in males aged 65-70 carriers of the rs877087 T allele having the bottom 20% of polygenic scores of body fat mass, lean mass and lipoprotein A was 1.28 (95% CI 0.96 to 1.7), whereas the relative risk was 2.9 (95% CI 1.38 to 6.11) for those having the top 20% of polygenic scores, compared to minimum risk group. In females aged 65-70 carriers of the rs877087 T allele, the relative risk increases to 4.4 (1.55 to 12.45) from 1.27 (0.85 to 1.91) with the highest polygenic scores (See supplementary table3 and Figure 3).

**Figure 3.**
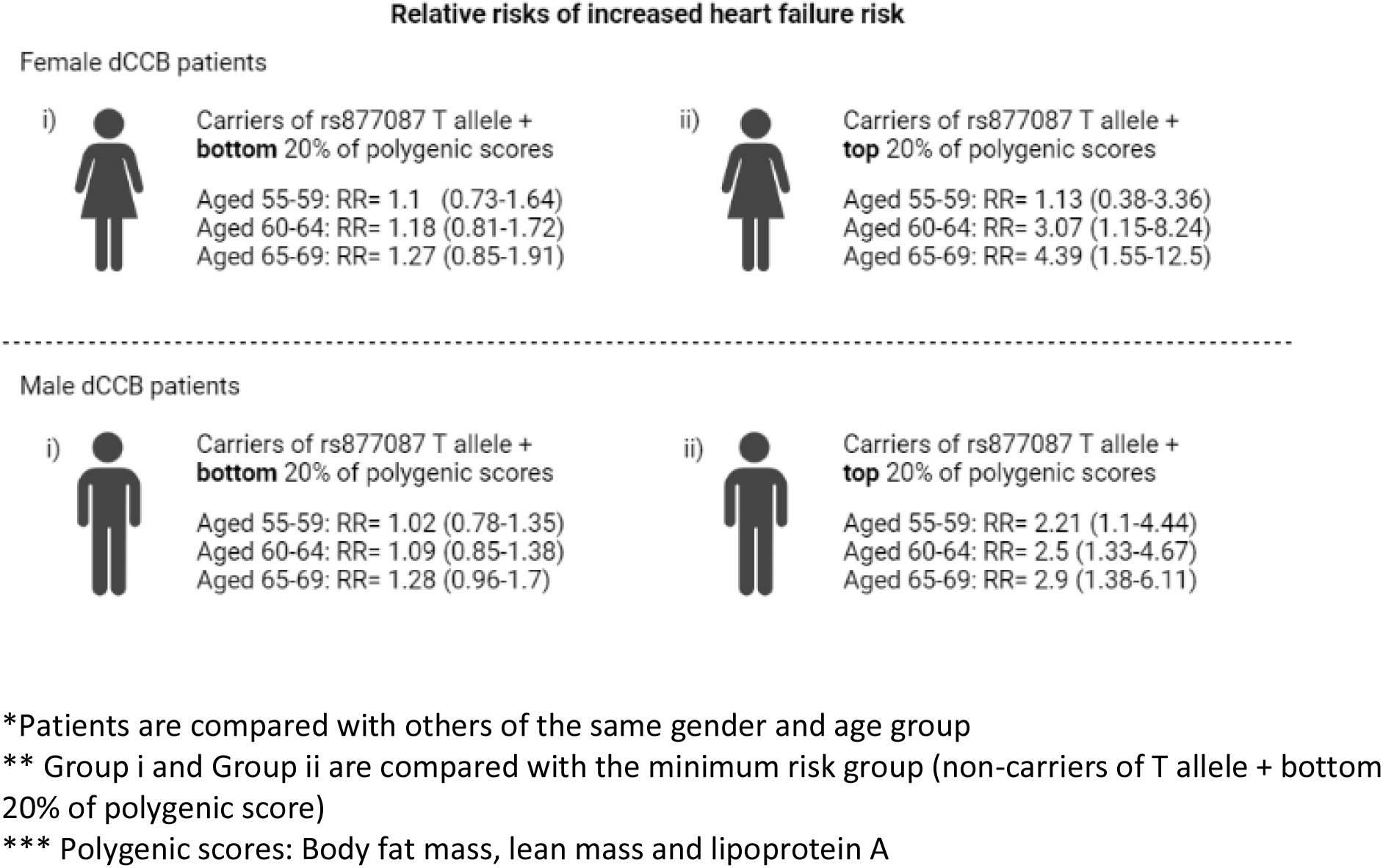
Polygenic score contribution on the risk of heart failure across age groups and sex in patients carrying rs877087 T allele.

#### Discontinuation

The relative risk of likelihood of discontinuation in males aged 60-65 carriers of rs10898815 A allele having the bottom 20% of polygenic scores of body fat mass and lean mass was 1.18 (95% CI 1.04 to 1.34), the risk increased to 1.51 (95% CI 1.16 to 1.96) for those having the top 20% of polygenic scores. The differences were more modest in women (See supplementary table 4 for details).

#### Sensitivity Analyses

1. In TwoSampleMR methods the weighted median and MR-Egger central estimates were consistent with those from IVW. In addition, we observed no significant evidence for bias due to unbalanced pleiotropy: MR-Egger intercept p=0.7 for heart failure, p=0.8 for discontinuation and p=0.2 for CHD (See supplementary table 5). The 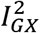 statistic for body fat mass exposures = 0.98, indicating no violation of the NOME assumption, thus MR Egger estimates were valid.
2. After excluding existing heart failure diagnoses, 1,737 heart failure diagnoses remained (N total=30,819). The polygenic scores increasing the risk of heart failure remained significant with similar effect sizes (i.e. HR per SD increase in polygenic score of body fat mass 1.13, 95% CI 1.07-1.18, *p*=1.08*10^**-6**^, of lipoprotein A HR 1.09, 95% CI 1.04-1.14, p=3.6*10^−4^).

## 5 Discussion

We previously reported that 5 pharmacogenetic variants (of 23 tested) were associated with CCB outcomes. Two variants (rs877087 T allele in gene RYR3, rs1089881 A allele in NUMA16) were more striking for heart failure and treatment discontinuation respectively, although the effect sizes were modest. A wide variety of individual patient characteristics have been reported as potentially modifying CCB responses. Here, we show that combining previously reported pharmacogenetic information with additional genetic data influencing patient characteristics - especially genetically predicted body fat mass - improves prediction of adverse clinical outcomes in hypertensive patients treated with dihydropiridine CCBs. By analyzing genetically predicted variation in individual characteristics we aimed to minimize confounding, as genetic variants are inherited at conception and are unchanged by later exposures or the downstream effects of disease. We provide the first evidence from a large-scale study of 32,000 dCCB patients combining polygenic scores with pharmacogenetic information reported. Previous genetic research in hypertension has modelled pharmacogenetic variants ^34^ or polygenic scores ^25^, but not together.

Adiposity is a common risk factor for many diseases, specifically with cardiovascular diseases^35–37^. Increasing in adiposity was found to increase the risk of cardiovascular diseases in UKB participants in a Mendelian randomization analysis^36^, another cross-sectional polygenic score study found an association between obesity and cardio-metabolic risk^35^. While our results support the current literature, we extend it by exploring associations in pharmacogenetics. We found that patients with genetically high burden of body fat mass were more likely to have heart failure and coronary heart disease despite the dCCB treatment, and increased risk of chronic kidney disease and discontinuation compared to those with low genetic burden. The pathway between higher body fat mass and clinical outcomes is likely two-fold: 1) via non-pharmacogenetic effects on cardio metabolic pathway, and 2) dCCBs are highly lipophilic, therefore greater body fat mass reduces medication effectives. Patients with higher fat mass may need larger doses^7,8^.

Genetically predicted lipoprotein A was found to have a causal effect on atherosclerotic cardiovascular diseases in several Mendelian Randomisation studies^38–40^. Although dihydropiridine CCBs were reported as being protective for experimental atherosclerosis ^41,42^, our results show that genetically predicted lipoprotein A were associated with increased risk of heart failure (HR 1.09: 95% CI 1.04-1.14) and coronary heart disease (HR 1.11: 95% CI 1.08-1.14) in dCCB patients. We were unable to test the associations with direct measured lipoprotein A at treatment initiation, however others have demonstrated similar predictions on cardiovascular diseases between the polygenic score and a direct measurement ^43^.

Analyses of data from a randomized controlled trial (RCT) ^10^ found that age and baseline BP were associated with CCB response in ∼60 patients. Also, a UK Biobank study that included patients who reported antihypertensive use at baseline found SBP polygenic score was associated with uncontrolled BP (Odds ratio: 1.70; 95% CI: 1.6-1.8)^27^, top vs. bottom quintile. Another UK Biobank study testing cardiovascular associations with SBP polygenic score at baseline found high SBP increased the cardiovascular outcomes in untreated hypertension with 1.04-fold and in treated hypertension with 1.06-fold^44^. In our study, high SBP and DBP polygenic scores are associated with decrease the risk for discontinuation in patients with high polygenic score (HR_SBP_: 0.93, 0.88-0.99 and HR_DBP_: 0.93, 0.87-0.99, top vs. bottom quintile), and high polygenic scores of DBP decrease the risk of heart failure and coronary heart disease in people treated with dCCB (HR_HF_: 0.86, 0.74-0.99 and HR_CHD_: 0.91, 0.84-0.99, top vs. bottom quintile) suggesting that those with the most severely increased blood pressures are more likely to continue treatment compared to those with more moderately increased pressures (although we lack data to test this directly). It might also suggest that patients with higher predisposition to hypertension may get better overall benefit from the CCB treatment, similar to the finding of an analysis of two RCT on lipid lowering showing that patients with highest genetic risk of atherosclerosis had better benefit from lipid lowering treatment^45^.

Serum calcium and urinary sodium were negatively associated with BP response in ∼52 Finnish men in another 4-week RCT ^12^, however we found no associations with serum calcium and sodium polygenic scores between outcomes in 32,000 patients. Calcium channel blockers have hepatic metabolism and renal excretion. Therefore, we tested the associations whether genetically predicted EGFR and fibrosis score affect CCB response. Results showed that high eGFR polygenic score increases the risk of chronic kidney disease in dCCB patients, but fibrosis polygenic score had no impact on any outcomes.

Dihydropiridine CCBs are accepted as safe options for the first line treatment of hypertension in non-black patients aged 55 and over, and reported to have protective effects in heart failure by the UK National Institute for Health and Care Excellence^32^. However, our results suggest that specific patients at higher genetic risk for heart failure (i.e., those with higher genetically predicted body fat mass, lean mass, and lipoprotein A, and carrying the pharmacogenetic variant rs877087 T allele) have worse outcomes compared to those with lower genetic risk. Individual level predictions incorporating this information might improve clinical outcomes in hypertension treatment. For example, while the risk ratio of heart failure in female aged 55-59 T allele carriers with high polygenic score of risk factors for heart failure is 1.13, it is 4.39 for those aged 65-69 compared to age and sex-matched patients with low risk.

Including genetic and non-genetic risk factors that could alter treatment response in routine prescribing could improve clinical outcomes. Individual patients’ characteristics such as biomarkers, weight, lipid and blood pressure levels would be easily accessible to clinicians. Under the evidence-based medicine regimes, more robust studies are required to implement new prescribing methods. In this study, using polygenic scores we minimize the effect of unmeasured confounders as genetic variants are fixed at conception and reflect lifetime exposure to the risk factor^21^. Current clinical utility of polygenic scores is not yet realized^46^, especially in pharmacogenetics due to smaller subgroup sample sizes^47^. We believe this is the first study examining polygenic scores of patients’ characteristics, and combining them with pharmacogenetic variants in a large 32,000 community patients prescribed calcium channel blockers with a long follow up over 5.9 years, using the primary care linked data reflecting the routine clinic. Studies such as these will inform efforts in personalized medicine.

This study is not without limitations: we were unable to test actual measurements of risk factors (routine GP data does not systematically measure these at treatment initiation). We were unable to analyze blood pressure response due to sparsity of blood pressure data available: only a subset of patients had GP-measured blood pressure in the primary care record within 2 months of treatment initiation. We were not able to analyze ‘dose’ due to lots of missing/inconsistent data, and high intra-patient variability. We were also unable to make specific recommendations for the high-risk group (e.g., should the dose be adjusted, or alternative treatment prescribed). Future studies are planned to extend the data analysis to incorporate untreated individuals and extend analytical methods – such as our recently published TWIST framework ^48^.

In summary, clinical outcomes seen in patients prescribed the common antihypertensives dCCB were better predicted when incorporating polygenic risk scores related to adiposity and lipoprotein A together with pharmacogenetic variants. Hence, efforts to personalize treatment regimes in CCB antihypertensive treatment should consider multiple genetic risk factors to improve patient outcomes. Combining pharmacogenetic and polygenic score data may have wider applications for optimizing prescribing of other medications, especially as genome wide genotype data becomes more widely available in routine clinical practice.

## Supporting information

dCCB modifiers supplementary tables

dCCB modifiers supplementary information

## Data Availability

The genetic and phenotypic UK Biobank data are available upon application to the UK Biobank (www.ukbiobank.ac.uk/register-apply).

## Funding and acknowledgements

DT is funded by the Ministry of National Education, Republic of Turkey. CP and DM are supported by the University of Exeter Medical School. JB is funded by an Expanding Excellence in England (E3) research grant awarded to the University of Exeter. JM is funded by a National Institute for Health Research Fellowship (NIHR301445). JD is also supported by the Alzheimer’s Society [grant: 338 (AS-JF-16b-007)]. This publication presents independent research funded by the National Institute for Health Research (NIHR). The views expressed are those of the author(s) and not necessarily those of the NHS, the NIHR or the Department of Health and Social Care. The funders had no input in the study design; in the collection, analysis, and interpretation of data; in the writing of the report; or in the decision to submit the article for publication. The researchers acted independently from the study sponsors in all aspects of this study.

Access to UK Biobank resource was granted under Application Number 14631. We would like to thank UK Biobank participants and coordinators for this dataset. The authors would like to acknowledge the use of the University of Exeter High-Performance Computing (HPC) facility in carrying out this work.

## Declaration of interest

Jack Bowden is employed part-time by Novo Nordisk, unrelated to the work presented here.

## Notes

### Competing Interest Statement

The authors have declared no competing interest.

### Author Declarations

The Northwest Multi-Centre Research Ethics Committee approved the collection and use of UK Biobank data (Research Ethics Committee reference 11/NW/0382).

