## Supplementary material for "Combining pharmacogenetics and patient characteristic polygenic scores to improve outcome prediction for Calcium Channel Blocker treatment": dCCB modifiers supplementary information

Türkmen et al.

**Supplementary Information**

**Figures**

- Figure 1- Heart failure rates across rs877087 T allele carriers compared to non-carriers of the allele
- Figure 2- Discontinuation rates across rs10898815 A allele carriers compared to non-carriers of the allele

**IVW MR Scatter Plots**

- Figure 3-MR of body fat mass and heart failure in UK Biobank dCCB prescribed patients
- Figure 4-MR of body fat mass and discontinuation in UK Biobank dCCB prescribed patients
- Figure 5-MR of body fat mass and coronary heart disease in UK Biobank dCCB prescribed patients

**Figure 1- Heart failure rates across rs877087 T allele carriers compared to non-carriers of the allele**

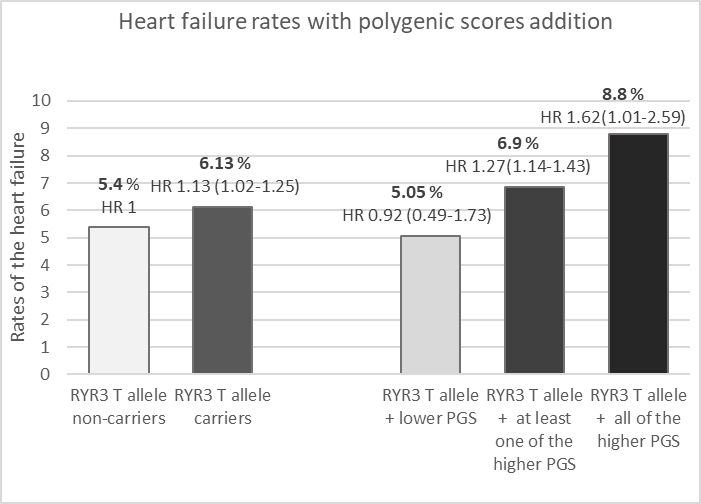

Only pharmacogenetic information assessed Pharmacogenetic and patients’ characteristics assessed together

*Lower PGS= Bottom 20% of polygenic scores of body fat mass, lean mass and lipoprotein A

*Higher PGS= Top 20% of polygenic scores of body fat mass, lean mass and lipoprotein A

**Figure 2- Discontinuation rates across rs10898815 A allele carriers compared to non-carriers**

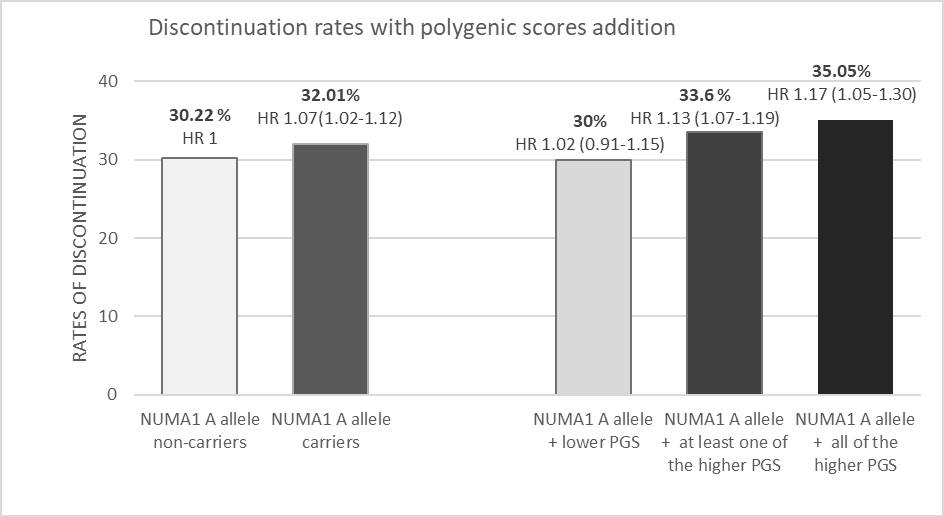

Only pharmacogenetic information assessed Pharmacogenetic and patients’ characteristics assessed together

*Lower PGS= Bottom 20% of polygenic scores of body fat mass and lean mass

*Higher PGS= Top 20% of polygenic scores of body fat mass and lean mass

**IVW MR Scatter Plots**

**Figure3-MR of body fat mass and heart failure in UK Biobank dCCB prescribed patients**

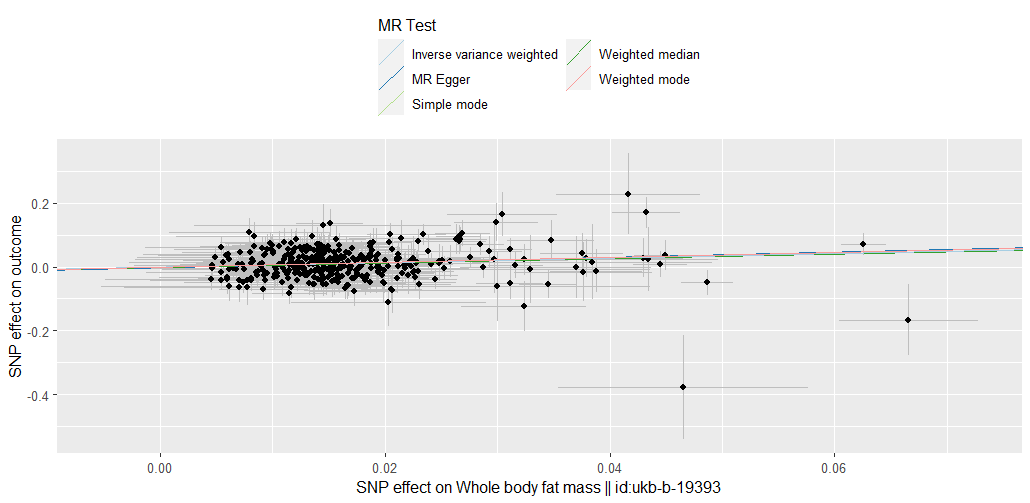

**Figure 4- MR of body fat mass and discontinuation in UK Biobank dCCB prescribed patients**

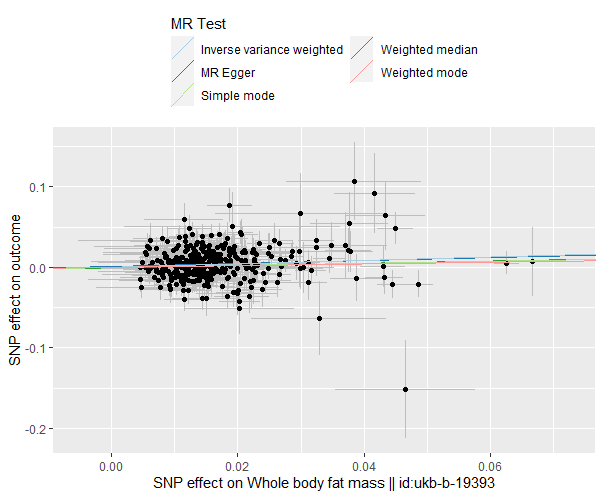

**Figure 5-MR of body fat mass and coronary heart disease in UK Biobank dCCB prescribed patients**

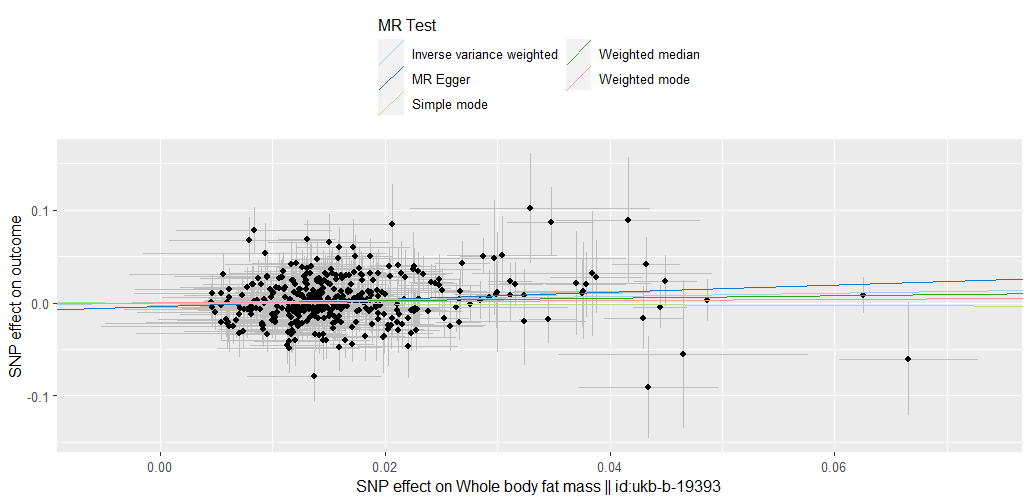
